# Digital Speech Analysis in Progressive Supranuclear Palsy and Corticobasal Syndromes

**DOI:** 10.1101/2020.09.18.20197657

**Authors:** Natalia Parjane, Sunghye Cho, Sharon Ash, Sanjana Shellikeri, Mark Liberman, Leslie M. Shaw, David J. Irwin, Murray Grossman, Naomi Nevler

## Abstract

**Background:** Progressive supranuclear palsy and corticobasal syndromes (PSPS-CBS) as well as nonfluent/agrammatic primary progressive aphasia (naPPA) are often due to misfolded 4-repeat Tau, but the diversity of the associated speech disorders beyond Apraxia of Speech (AoS) is poorly understood.

**Objective:** Investigate the full range of acoustic and lexical properties of speech to test the hypothesis that PSPS-CBS show a subset of speech impairments found in naPPA.

**Methods:** Acoustic and lexical measures, extracted from natural, digitized semi-structured speech samples using novel, automated methods, were compared in PSPS-CBS (n=87), naPPA (n=25) and healthy controls (HC, n=41). We also explored speech in a group of PSPS-CBS patients with concomitant naPPA (PSPS-CBS+naPPA, n=8). We related these measures to grammatical performance and speech fluency, core features of naPPA, and to cerebrospinal fluid (CSF) phosphorylated tau (pTau) in patients with available biofluid.

**Results:** Both naPPA and PSPS-CBS speech had shorter speech segments, longer pauses, higher pause rate, reduced fundamental frequency (f0) range, and slower speech rate compared to HC. naPPA speech was distinct from PSPS-CBS with shorter speech segments, more frequent pauses, slower speech rate, and reduced verb production. In both groups, acoustic duration measures generally correlated with speech fluency and grammatical performance. PSPS-CBS+naPPA resembled naPPA in most speech measures and had the narrowest f0 range. CSF pTau levels correlated with f0 range and verb production in PSPS-CBS and naPPA.

**Conclusion:** The speech pattern of PSPS-CBS overlaps that of naPPA apart from AoS, and may be related to CSF pTau.

## INTRODUCTION

Progressive supranuclear palsy syndrome and corticobasal syndrome (PSPS-CBS) are both frontotemporal degeneration (FTD) spectrum disorders commonly associated with underlying 4-repeat tau pathology with mean age of onset in the 60s and 7-year life expectancy. PSPS-CBS are noted for prominent extrapyramidal motor symptoms and higher cognitive dysfunction, with MRI cerebral atrophy and glucose-PET hypometabolism in subcortical structures and frontal cortex, sometimes extending to temporal and parietal areas [1-4].

While researchers often focus on motor symptoms in PSPS-CBS, cognitive symptoms are increasingly recognized. These symptoms may include deficits in executive, visuo-spatial, memory, social, and language functions. Speech abnormalities have focused on apraxia of speech (AoS), but can also include difficulty with naming, word finding, and phonological expression as well as dysfluent, slow, slurred, and hypophonic speech [5-9]. Sometimes patients present with concomitant nonfluent/agrammatic variant of primary progressive aphasia (naPPA) [1-3].

naPPA, an FTD disorder which also frequently has underlying tau pathology [10,11], is characterized by AoS, but also includes slow, dysfluent, effortful speech, agrammatism, speech-sound errors, and reduced pitch range and verb production [12-19]. Rare case studies and small series reporting PSPS-CBS patients with concomitant naPPA show that patients can have severe speech impairments, like those found in naPPA. Such patients can either present with PSPS-CBS or naPPA at onset, and motor and cognitive symptoms vary depending on the primary presentation [20-31].

In the context of these observations, there is an on-going debate regarding the source of speech and language impairments in PSPS-CBS, attributing these to either cognitive or motor mechanisms. Many studies focus on motor speech abnormalities such as dysarthria and AoS, but do not investigate potential cognitive effects on speech. Studies directly comparing PSPS-CBS and naPPA would be important to improve our understanding of the underlying mechanisms of speech impairment. However, studies directly comparing speech and language impairments in these disorders are rare and often rely on neuropsychological tests and subjective assessments [25,28,30,31,32].

This study aims to characterize speech and language in PSPS-CBS compared to healthy controls (HC) and naPPA and to relate speech markers to underlying cognitive mechanisms and cerebrospinal fluid (CSF) levels of phosphorylated tau (pTau), thought to be a marker of underlying neuropathology [33]. Utilizing novel automated techniques to analyze speech from a picture description task, we quantified speech markers, including acoustic measures such as pitch, speech and pause segment durations, and pause rate, as well as lexical measures such as partial word, verb and noun productions. We hypothesized that acoustic and lexical properties of speech in PSPS-CBS would form a spectrum with naPPA. PSPS-CBS was thus expected to exhibit higher pause rate and reduced speech segment duration, pitch range, and speech rate, reduced verb production, and produce more partial words than healthy controls. We also directly compared PSPS-CBS with naPPA, and separately examined a subgroup of PSPS-CBS patients with concomitant naPPA (PSPS-CBS+naPPA). We further hypothesized these speech markers would correlate with CSF pTau levels.

## METHODS

### Study Population

We examined 153 native English speakers with PSPS (n=41), CBS (n=46), and naPPA (n=25), and healthy controls (HC, n=41). Patients were examined at the Hospital of the University of Pennsylvania by experienced neurologists (MG, DJI) and the clinical phenotype was reviewed in a multidisciplinary consensus conference according to published criteria [1,3,15,34]. All patients were assessed between January 2000 and June 2019. Some of the naPPA patients have been reported previously, but not in comparison to PSPS-CBS [12-14,17,18,35]. We excluded cases with other neurological (e.g. vascular disease, hydrocephalus, head trauma), medical (e.g. infectious, inflammatory, metabolic) or primary psychiatric (e.g. psychosis, major depression, bipolar) conditions that could confound our observations. We excluded 4 patients with PSPS-CBS diagnosis because it was not clear whether the primary diagnosis was PSPS or CBS. Demographic and clinical characteristics of the patient groups are summarized in Table 1. We combined patients with PSPS and CBS who did not have concomitant naPPA (PSPS-CBS, n=87) since these groups were matched for sex, age, education, and disease duration, and their speech characteristics were similar (Supplementary Table 1). However, since MMSE scores differed in PSPS and CBS patients (*t=*2.71, *p=*0.009, 95% confidence interval (CI) 0.74 to 4.88), we also compared PSPS and CBS groups separately for speech and language impairments (see supplementary material). PSPS-CBS and naPPA also were matched for education, sex, age, and disease duration. Evaluation of MMSE scores with Tukey pairwise comparisons showed that PSPS-CBS (*p<*.001) and naPPA (*p<*.001) were impaired compared to HC (*F*=16.32, 95% CI 0.08 to 0.31), but the patient groups did not differ from each other.

**Table 1:** Demographic and clinical characteristics of patient groups and healthy controls.

| Mean (Standard Deviation) | HC | PSPS-CBS | naPPA | PSPS-CBS+naPPA | *P*-value<sup>1</sup> |
| --- | --- | --- | --- | --- | --- |
| **n** | **41** | **87** | **25** | **8** |  |
| **Age (y)** | 68.94 (7.57) | 68.16 (7.53) | 70.91 (8.93) | 71.20 (6.15) | **0.298** |
| **Education (y)** | 15.80 (2.42) | 14.75 (2.69) | 15.32 (3.00) | 15.75 (1.98) | **0.108** |
| **Disease Duration (y)** | NA | 4.13 (2.42)<sup>3</sup> | 3.32 (1.91) | 3.00 (1.07) | **0.128** |
| **Sex: M (%)** | 18 (43.9%) | 40 (46%) | 11 (44%) | 4 (50%) | **0.969** |
| **n** | **35** | **74** | **24** | **7** |  |
| **MMSE<sup>2</sup> (max = 30)** | 29.11 (0.99) | 24.92 (4.75)* | 23.04 (5.89)* | 27.14 (2.41) | **<0.001** |

Abbreviations: y = year; MMSE = Mini-Mental State Examination; M = males.
<sup>1</sup>p-values for all contrasts in main groups: HC, PSPS-CBS, and naPPA
\*differs from HC ( $p < 0.001$ )
<sup>2</sup>MMSE obtained within 6 months of speech sample
<sup>3</sup>n=86 PSPS-CBS patients

To investigate how a combination of underlying motor and cognitive impairment can affect speech and language patterns, we explored speech characteristics in a small, separate group of patients with PSPS-CBS and concomitant naPPA (PSPS-CBS+naPPA, n=8), who were not included in the PSPS-CBS or naPPA patient groups. We excluded one patient with CBS, naPPA, and concomitant behavioral variant frontotemporal degeneration to constrain the sub-analysis to PSPS-CBS with a concomitant naPPA diagnosis. PSPS-CBS+naPPA were matched demographically with both patient groups.

This study complies with guidelines on human experimentation. All participants were enrolled in study protocols and participated in an informed consent procedure in accordance with the Declaration of Helsinki approved by the Institutional Review Board of the University of Pennsylvania. The appropriate institutional forms have been archived.

### Experimental speech task

We used the same methodology to acquire a semi-structured speech sample as employed in our previous studies [13,14,18,36,37,42]. Briefly, an interviewer instructed participants to describe the Cookie Theft scene from the Boston Diagnostic Aphasia Examination [38] in as much detail as they could and using full sentences. The interviewer prompted the participants to continue speaking if they were silent for more than 10 s, while aiming to minimize interruptions and speech overlap with participant’s speech. Total recording mean duration was 70.9 s (range 15.1-156.6 s). Longitudinal recordings were excluded, and only the earliest recordings of participants were analyzed. Recordings were obtained digitally in a quiet room with minimal background noise and stored as .wav files.

### Acoustic analysis

We used the same methodology as published previously to analyze the digitized speech samples [18,36,37,42]. Briefly, a speech activity detector (SAD), developed at the University of Pennsylvania Linguistic Data Consortium (LDC), automatically segmented the acoustic signal into speech and non-speech (silent pause) segments [39,40]. The output was manually reviewed in Praat [41] to validate the accuracy of the automated algorithm and corrections were sparse. We manually labelled out interviewer’s speech as well as cough, laughter, and background noise that interrupted speech in order to minimize pitch tracking errors.

Using an R script, we extracted and calculated acoustic measures, including mean speech segment duration, mean pause segment duration, and pause rate (number of pauses per minute over total speaking time) from the SAD, and excluded silences at the beginning and end of recordings. We tracked the fundamental frequency (f0) range of the participant using Praat’s pitch tracker. Pitch tracking provided f0 estimates for each 10 milliseconds in Hertz during continuous speech segments. We transformed f0 range to the semitone (ST) scale, a relative logarithmic scale and focused on the participants’ trimmed f0 range, which represents pitch range as described in our previous studies [18,36,37,42]. We reviewed all participants with a pitch range 1.5 SD above or below the group mean and found 23 participants with substantial “creaky voice” or substantial background noise. These vocal characteristics carry a high probability for pitch-tracking errors, so we excluded these participants from further analyses of f0 range (HC, n=5; PSPS, n=5; CBS, n=8; naPPA, n=5; PSPS-CBS+naPPA, n=1).

### Lexical semantic analyses

We used the same methodology as previously reported to perform an analysis of part of speech (POS) [35,42] to automatically tag POS of the Cookie Theft descriptions [43,44]. Briefly, spaCy [44] uses the Penn Treebank [45] to map words onto tags from the Google Universal POS tag set [46]. Characteristics of naPPA [15] include non-fluent speech, speech errors, and grammatical errors and simplifications, and we analyzed patients’ speech for these characteristics. Speech rate (words per minute, wpm), a measure of speech fluency, was calculated automatically by adding up word counts for each recording from the spaCy output, then dividing by total recording time, excluding interviewer speech. Word categories included adjective, preposition, adverb, coordinating conjunction, determiner, filler, noun, numeral, particle, pronoun, proper noun, verb, partial word, as well as tokens that could not be identified by the language model. These tokens were annotated as “X,” and upon manual inspection consisted mostly of partial words and thus named “partial words” in our analyses. We also analyzed verb use because of the close relationship between this POS and grammatical expression, and we selected nouns as a control category that is less closely related to grammatical expression. In our analyses, instances of partial words, verbs, and nouns per 100 words were calculated to control for speech sample size.

We previously performed manual coding for measures of “grammatical performance,” reflecting the grammatical complexity of participants’ utterances [13]. We selected two such grammatical measures that were previously linked to naPPA speech [12,13,14,18]: dependent clauses per 100 utterances (DC) and percentage of utterances that are well-formed sentences (WFS). We calculated the average of the values, in order to minimize ceiling and floor effects.

These measures were used in the clinical correlations described below to validate our novel automated speech markers.

### Cerebrospinal fluid phosphorylated tau analysis

We analyzed CSF samples that were available in a subset of patients, including PSPS-CBS (n=50; PSPS*=*26 and CBS=24), naPPA (n=11), and PSPS-CBS+naPPA (n=5). As previously reported [33], CSF was analyzed using two platforms - Luminex xMAP or Innotest ELISA (which is then transformed to the Luminex scale). We previously associated CSF levels of pTau directly with pathologic burden of tau in our autopsy cohort [33]. Using a pathologically validated algorithm, we screened patients for possible underlying AD pathology with the ratio pTau/Aβ<0.09 [33]. This screening left us with 47 patients (PSPS-CBS, n=34, including PSPS*=*21 cases and CBS=13; naPPA, n=9; PSPS-CBS+naPPA, n=4) whose CSF profiles suggest underlying FTLD pathology.

### Statistical analysis

All statistical analyses were conducted using R v.3.6.3, RStudio v.1.2.5033 [47]. We evaluated normal distribution of speech variables using Kernel density and Q-Q plots. Levene’s test revealed violations of homogeneity of variances for pause segment duration, pause rate, partial words, and nouns, so we used non-parametric tests, including the Kruskal-Wallis test to compare groups on acoustic, lexical, and grammatical performance. Pairwise comparisons between groups were calculated with the Mann-Whitney test with a 95% CI. For comparisons between groups, unless otherwise stated, all differences are significant at least at the two-tailed level *p<*0.05 and the p-value was adjusted using a False Discovery Rate (FDR) correction for multiple comparisons. Since speech rate included two PSPS-CBS outliers (at 232 wpm and 289 wpm), we checked to see if removing these outliers had leverage on our results. They had no leverage on regression results, so we report results including these outliers. In RStudio, additional packages were used [48-54].

To investigate the relationship between acoustic-lexical markers and cognitive deficits and because naPPA is characterized by grammatical simplification and slowed speech [15], we performed linear regression analyses where each acoustic-lexical marker was the outcome (dependent) measure, and grammatical performance and speech rate (each was used in a separate model) were the main predictors (independent variable) in a cohort that included both main patient groups (PSPS-CBS and naPPA). We first examined group interactions with grammatical performance or speech rate (eg. [speech marker] ∼ [grammatical score] + [group] + [grammatical performance X group]). We eliminated the interaction term when no interaction was found. We used natural logarithmic transformations to normalize skewed data when examining speech and pause segment durations in regressions, relating these markers to grammatical performance and speech rate. Regression models were validated by viewing the residual plots.

To link the speech markers with probable underlying pathology, we used linear regression models where CSF pTau level (transformed to the natural log) was the dependent variable. We ran a separate model for each speech marker as predictor (independent variable). We controlled for potential confounders including age and the time interval between the speech sample collection and CSF collection (−62.7-106.7 weeks). We checked residual plots to validate the models.

In an exploratory analysis exploring speech markers in a group with combined PSPS-CBS+naPPA, we compared their speech markers to those of the main patient groups, using the Mann-Whitney test. Due to the small sample size the results of this exploratory analysis were not robust for corrections and thus we report uncorrected p-values.

## Data Availability

Access to all data anonymized for purposes of replicating procedures and results is available upon request. We share anonymized data with qualified investigators who have appropriate regulatory approval and transfer agreements.

## RESULTS

### Acoustic results

Table 2 summarizes acoustic, lexical, and grammatical characteristics of participants. Pairwise comparisons of acoustic markers revealed that, compared to HC, PSPS-CBS had shorter speech segments (W=722, *p<*.001, 95% CI −0.77 to −0.4, Fig. 1A), longer pause segments (W=2914, *p<*.001, 95% CI 0.46 to 0.98, Fig. 1B), higher pause rate (W=2814, *p<*.001, 95% CI 9.29 to 18.15, Fig. 1C), and reduced f0 range (W=975, *p=*.03, 95% CI −1.84 to −0.18, Fig. 1D).

**Table 2:** Acoustic, lexical, and grammatical characteristics of patient groups and healthy controls.

| Mean (Standard Deviation) | HC | PSPS-CBS | naPPA | PSPS-CBS+naPPA | P-value<sup>1</sup> |
| --- | --- | --- | --- | --- | --- |
| **n** | **41** | **87** | **25** | **8** |  |
| **Speech Segment Duration (s)** | 2.00 (0.57) | 1.42 (0.48)<sup>*,P</sup> | 1.17 (0.50)<sup>*</sup> | 1.09 (0.35) | **<0.001** |
| **Pause Segment Duration (s)** | 0.93 (0.44) | 1.82 (1.01)<sup>*</sup> | 1.65 (1.10)<sup>*</sup> | 1.13 (0.41) | **<0.001** |
| **Pause Rate (ppm)** | 31.61 (9.51) | 45.10 (13.43)<sup>*,P</sup> | 57.81 (19.95)<sup>*</sup> | 59.77 (18.18) | **<0.001** |
| **n (f0 Range)** | **36** | **74** | **20** | **7** |  |
| **f0 Range (ST)** | 5.87 (1.94) | 4.97 (2.03)<sup>*</sup> | 4.30 (1.59)<sup>*</sup> | 3.56 (1.21) | **0.011** |
| **Speech Rate (wpm)** | 145.20 (32.31) | 92.31 (42.38)<sup>*,P</sup> | 65.26 (25.54)<sup>*</sup> | 70.40 (29.94) | **<0.001** |
| **Partial Words** | 0.54 (0.87) | 0.95 (1.47) | 3.48 (5.25)<sup>*</sup> | 1.12 (1.47) | **<0.001** |
| **Verb Production** | 23.03 (3.41) | 22.94 (4.47)<sup>P</sup> | 19.97 (4.48)<sup>*</sup> | 21.82 (3.27) | **0.006** |
| **Noun Production** | 20.21 (4.44) | 22.52 (5.19)<sup>*</sup> | 21.49 (8.38) | 22.12 (5.69) | **0.1** |
| **Grammatical Performance Score<sup>2</sup>** | -0.00 (0.78) | -0.84 (1.13)<sup>*</sup> | -1.48 (1.71)<sup>*</sup> | -1.34 (1.18) | **<0.001** |

Abbreviations: s = seconds; ppm = pauses per minute of speech time; ST = semitones; wpm = word per minute.
<sup>1</sup>p-values for all contrasts in main groups: HC, PSPS-CBS, and naPPA:
<sup>\*</sup>differs from HC
<sup>P</sup>differs from naPPA
<sup>2</sup>values are z-scored using control mean and standard deviation

**FIGURE 1.**
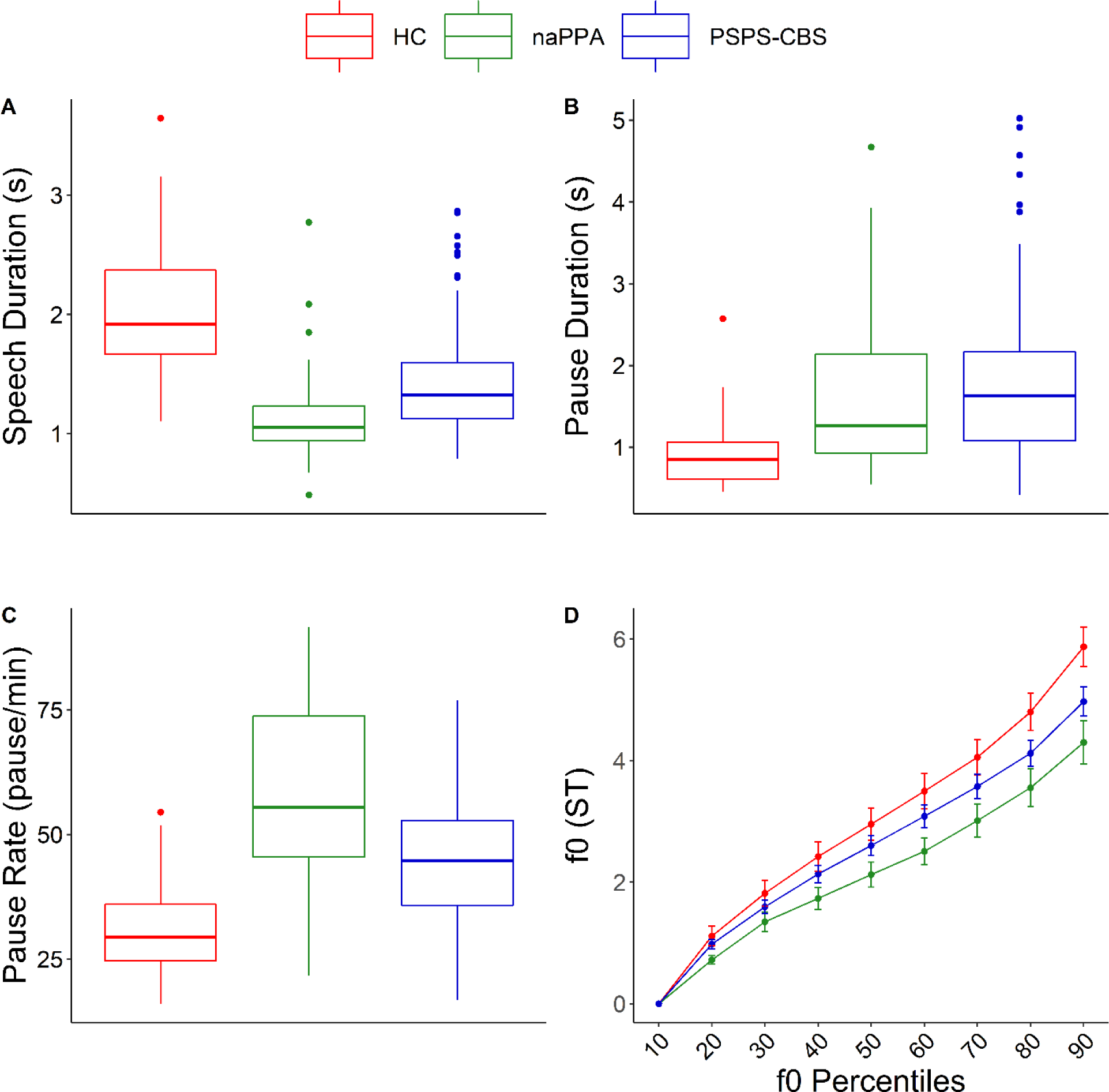
Speech and Pause durations and f0 range by clinical phenotype. (A) Mean speech segment duration measured in seconds. (B) Mean pause segment duration measured in seconds. (C) Pause rate measured in pauses per minute of speech time (ppm). (D) fundamental frequency (f0) range in semitones (ST): the 90^th^ percentile represents the trimmed f0 range. HC, healthy controls; naPPA, non-fluent/agrammatic primary progressive aphasia; PSPS-CBS, progressive supranuclear palsy and corticobasal syndrome spectrum disorders; s, seconds.

Compared to HC, naPPA also had shorter speech segments (W=110, *p<*.001, 95% CI - 1.1 to −0.59, Fig. 1A), longer pause segments (W=761, *p=*.001, 95% CI 0.18 to 0.74, Fig. 1B), higher pause rate (W=899, *p<*.001, 95% CI 17.93 to 33.38, Fig. 1C), and reduced f0 range (W=186, *p=*.007, 95% CI −2.62 to −0.62; Fig. 1D).

Compared to naPPA, PSPS-CBS patients produced longer speech segments (W=671, *p=*.004, 95% CI 0.09 to 0.41) and had reduced pause rate (W=1526, *p=*.002, 95% CI −20.42 to - 4.37).

An exploratory analysis of PSPS-CBS+naPPA revealed an acoustic pattern similar to that of naPPA. Like naPPA, PSPS-CBS+naPPA had shorter speech segments (W=200, *p=*.048, 95% CI −0.58 to −0.004) and higher pause rate compared to PSPS-CBS (W=504, *p=*.037, 95% CI 1.03 to 28.64). PSPS-CBS+naPPA also produced shorter pause segments (W=193, *p=*.038, 95% CI - 1.07 to -.03) and showed reduced f0 range compared to PSPS-CBS (W=144, *p=*.054, 95% CI - 2.7 to 0.02), but not naPPA (Supplementary Figure 1).

### Lexical results

Compared to HC, PSPS-CBS had a slower speech rate (W=486, *p<*.001, 95% CI −71.36 to −46.22, Fig. 2A) and produced more nouns (W=2282.5, *p=*.033, 95% CI 0.55 to 4.27, Fig. 2B). They produced a similar number of partial words and verbs (Fig. 2B).

**FIGURE 2.**
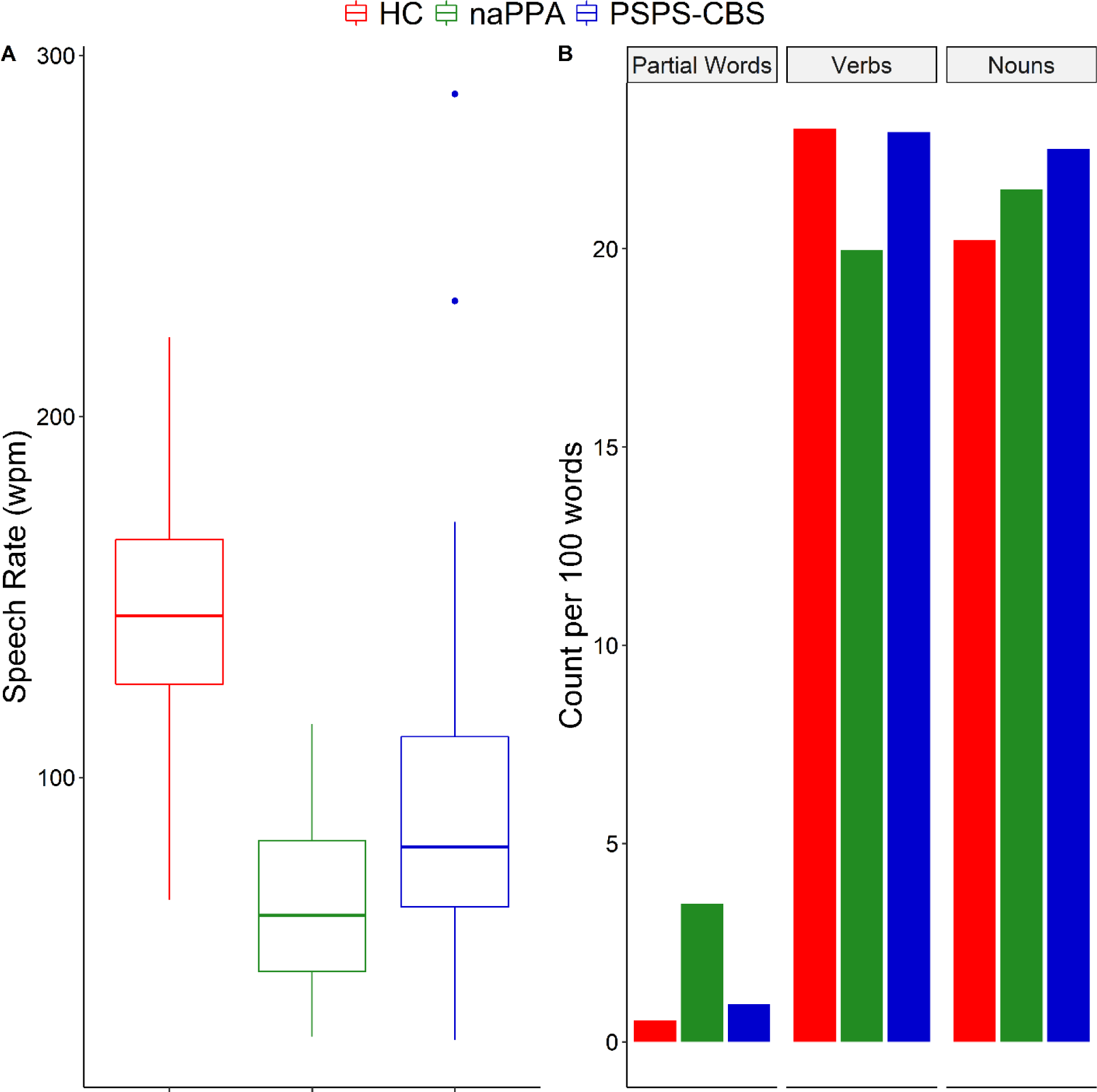
Lexical measures by clinical phenotype (A) Speech rate in words per minute (wpm). (B) Partial words, verb and noun production counts per 100 words. HC, healthy controls; naPPA, non-fluent/agrammatic primary progressive aphasia; PSPS-CBS, progressive supranuclear palsy and corticobasal syndrome spectrum disorders.

Compared to HC, naPPA also had a slower speech rate (W=30, *p<*.001, 95% CI −95.79 to −66.18, Fig. 2A). They produced fewer verbs than HC (W=318.5, *p=*.016, 95% CI −4.86 to −0.7, Fig. 2B) and marginally more partial words (W=678, *p=*.064, 95% CI 0 to 1.32, Fig. 2B). They did not differ from HC (*p=*.60) or PSPS-CBS (*p=*.54) in their noun production (Fig. 2B). naPPA also had a slower speech rate compared to PSPS-CBS (W=647, *p<*.002, 95% CI −35.43 to −7.95) and produced fewer verbs (W=717.5, *p=*.016, 95% CI −4.85 to −0.62).

In an exploratory analysis, we saw that PSPS-CBS+naPPA speakers had similar lexical markers and speech rate to the two other patient groups. naPPA seemed to have produced more partial words and fewer verbs, but this was not statistically significant (Supplementary Figure 2).

### Clinical Correlations

We used linear regression models to test the hypothesis that our digitized, automated, acoustic and lexical markers were associated with grammatical impairment and speech rate as these are the most distinctive linguistic manifestations of naPPA [15].

Grammatical impairment was associated with pause segments (beta=-1.35, *p<*.001, Fig. 3B), pause rate (beta=-21.73, *p=*.013, Fig. 3C), and verb production (beta=9.95, *p<*.001, Fig. 3D), independent of phenotype. For speech segments, this association was only found for PSPS-CBS (beta=0.69, *p=*.028, Fig. 3A). Grammatical impairment did not correlate with partial words or f0 range.

**FIGURE 3.**
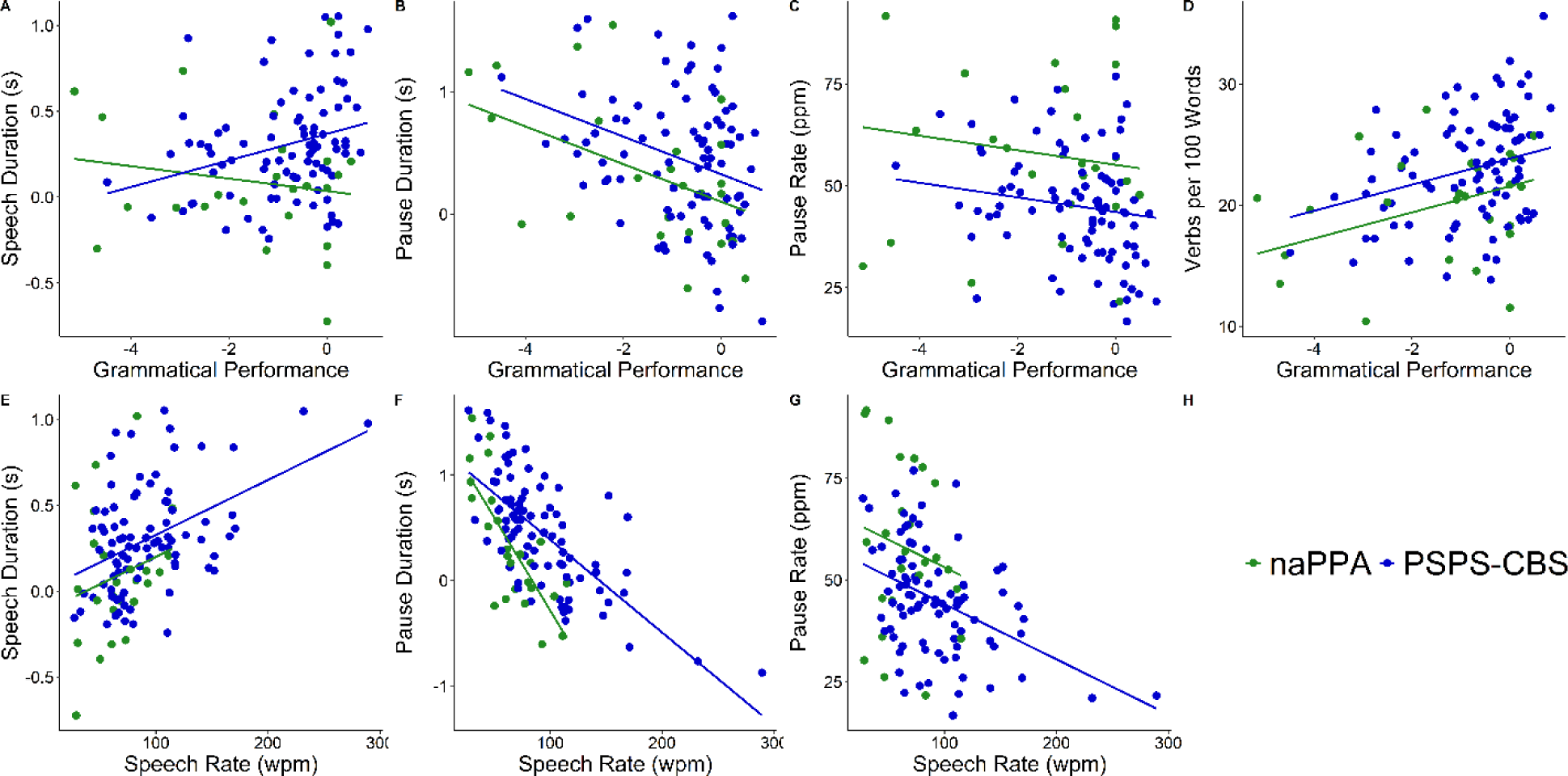
Linear regression models relating grammatical performance, here transformed by z-score relative to healthy controls, to (A) the natural log of mean speech segment duration measured in seconds, (B) natural log of mean pause segment duration measured in seconds, (C) pause rate measured in pauses per minute of speech time (ppm), (D) and verb count per 100 words, (E) and relating speech rate in words per minute (wpm) to the natural log of mean speech segment duration, (F) natural log of mean pause segment duration, and (G) pause rate. naPPA = non-fluent/agrammatic primary progressive aphasia; PSPS-CBS = progressive supranuclear palsy and corticobasal syndrome spectrum disorders; s = seconds.

Speech rate was found to be associated with speech segments (beta=0.003, *p<*.001, Fig. 3E) and pause rate (beta=-0.14, *p<*.001, Fig. 3G), independent of phenotype. Pause segments were related to speech rate, such that with slower speech, pause segments became longer, and this was more pronounced in naPPA (beta=-0.018, *p<*.001, Fig. 3F) than PSPS-CBS (beta=- 0.009, *p<*.001). Speech rate did not correlate with f0 range, partial words, verb or noun production. Speech rate did correlate with grammatical impairment (beta=104.58, *p<*.001), independent of phenotype.

There were no associations between the acoustic measures and partial words or verb production. Grammatical impairment correlated with noun production (beta=-13.741, *p<*.001), independent of phenotype. However, we found no association between nouns and speech segments (*p*=.55), pause segments (*p*=.26), pause rate (*p*=.85), or f0 range (*p*=.63).

### CSF analyses

We related CSF pTau levels in our patient groups to their acoustic-lexical properties using linear regression models. We found narrow f0 range (beta=-0.069, *p=*.002, Fig. 4A) and lower verb production (beta=-0.023, *p=*.034, Fig. 4B) to be related to higher levels of pTau in CSF of all patients.

**FIGURE 4.**
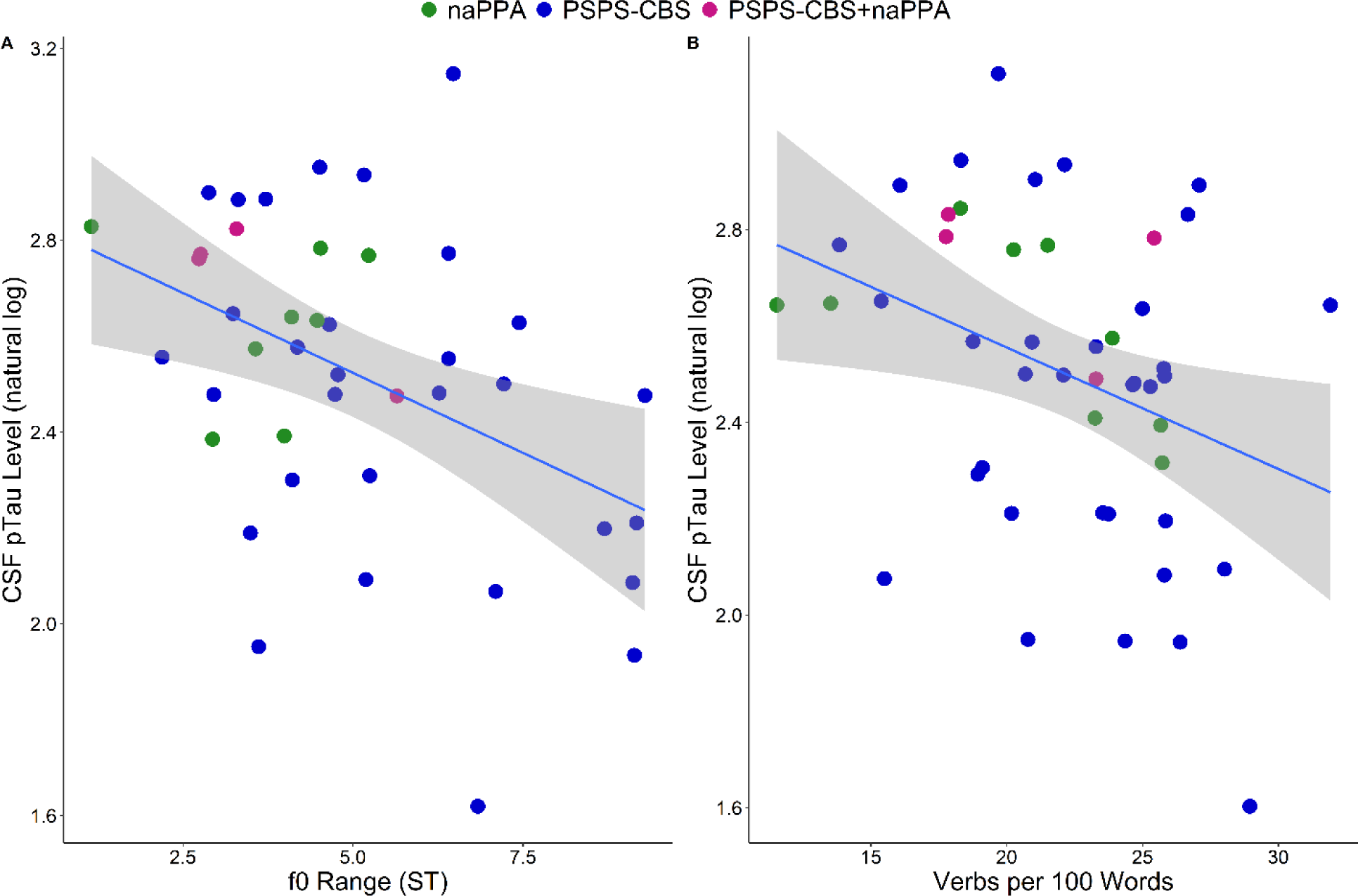
A linear regression model relating the natural logarithm of CSF pTau levels to (A) f0 range and (B) verb count per 100 words, while controlling for age and time interval between speech sample recording and CSF collection. CSF pTau = cerebrospinal fluid phosphorylated tau; naPPA = non-fluent/agrammatic primary progressive aphasia; PSPS-CBS = progressive supranuclear palsy and corticobasal syndrome spectrum disorders; PSPS-CBS+naPPA = PSPS-CBS with concomitant naPPA; f0 = fundamental frequency; ST = semitones.

## DISCUSSION

We used a novel, automated speech analysis of natural speech to characterize acoustic and lexical markers in PSPS-CBS and naPPA. We found that speakers with PSPS-CBS produce shorter speech segments than normal speakers, longer and more frequent pauses, as well as reduced f0 range and speech rate. naPPA speakers showed a similar pattern of acoustic markers, but their speech segments were even shorter and pause rate higher. Higher production of partial words and reduced verb production were only found in naPPA speakers. Longer and more frequent pauses were related to impaired grammaticality in both PSPS-CBS and naPPA, and shorter speech segments were related to impaired grammaticality in PSPS-CBS. Longer and more frequent pauses and shorter speech segments were related to slow speech rate in PSPS-CBS and naPPA. Narrow f0 range and reduced verb production were related to higher pTau levels in patients’ CSF. We will discuss these findings in detail below.

We found that PSPS-CBS and naPPA showed similar patterns of impairment in acoustic properties of speech. This is an important finding because of the claim that slow, dysfluent, and agrammatic speech found in naPPA [12-18] can be an early predictor of PSPS and CBS [21,55]. Our findings suggest that PSPS-CBS speakers indeed lie on a spectrum of speech impairments with naPPA, and that slowed speech may simply be a property of the PSPS-CBS phenotype. A previous study reported similar aprosodic speech and vocabulary in PSPS and naPPA [56]. PSPS speakers were also found to have reduced speech and pitch range, and a higher pause rate compared to Parkinson’s disease [57]. Our findings of acoustic impairment in PSPS are in line with these reports and we extend these findings to CBS.

While PSPS-CBS and naPPA exhibited some similar speech characteristics, it is important to highlight that they do not have identical speech patterns. naPPA speakers are more impaired in their acoustic markers and have distinct lexical impairments. Our findings are in keeping with previous publications indicating that naPPA is characterized by effortful, agrammatic speech, with reduced pitch range and verb production, higher pause rate and lower speech rate [12-18,35]. Importantly, our findings suggest that pause rate, speech rate, and lexical measures, including partial words and verbs, can be used as markers to distinguish between PSPS-CBS and naPPA.

It is unfortunate that we do not have an automated assessment of speech errors, due in part to the difficulty detecting and quantifying speech sounds that are not from the corpus of speech sounds in English, the native language of our speakers. It is important to be able to quantify speech errors since this represents an important characteristic of AoS [58]. However, partial words may in part be a surrogate for detecting AoS because this characteristic may be related in part to mid-word pauses that are another characteristic of AoS. Future longitudinal work is needed to determine whether the presence of shorter speech segments, increased pause rate, lower speech rate, increased partial words and reduced verb production distinguishes between those naPPA patients who progress to a PSPS-CBS phenotype and those who do not.

Our cohort of 8 participants with PSPS-CBS+naPPA is a small but informative group. We found that this group of speakers is generally similar to naPPA in their acoustic markers with shorter speech segments and higher pause rate compared with PSPS-CBS. They also exhibited a reduced f0 range compared to PSPS-CBS. Although we do not have longitudinal data in these cases, these observations support the hypothesis that impaired speech markers found in PSPS-CBS can become exacerbated to the point where these patients meet criteria for naPPA. These results align with earlier case studies on patients with concomitant PSPS-CBS and naPPA, where researchers subjectively observed that speech was more severely impaired than might be expected in PSPS-CBS alone [20-29].

Reduced grammatical complexity and dysfluency are said to be distinct characteristics of naPPA [12-18]. The results of our regression analyses support the hypothesis that longer pauses, shorter speech segments and higher pause rate relate to impaired grammaticality and dysfluency in patients with naPPA as well as patients with PSPS-CBS. Reduced speech rate and verb production were also related to grammatical impairment in both PSPS-CBS and naPPA. The selectivity of this finding is further supported by the fact we did not find an association between the same speech markers and noun production in any of the patient groups. These observations reinforce the assertion that speech in naPPA and PSPS-CBS can be considered to be on the same spectrum of impairment.

Finally, we found an association between CSF pTau levels and f0 range and verb production. This replicates our previous study where we found that CSF pTau is related to f0 range in naPPA patients [18]. Here, we extend this finding to PSPS-CBS, which is also a tauopathy. These findings indicate a link between our novel acoustic-lexical markers and underlying tau pathology. Thus, f0 range and verb production may be inexpensive markers that can screen for underlying tau pathology in PSPS-CBS and naPPA. Since speech is easily collected and is reliable and reproducible, this brings out a potential role for these acoustic-lexical markers in patient screening and monitoring during clinical trials for novel interventions targeting tau pathology.

### Strengths and limitations

The automated speech analyses described in this study are reliable and objective. No subjective ratings were used which could confound the data. The SAD is accurate in extracting pause and speech measures from the natural speech of patients, and the POS tagger has a high accuracy in tagging parts of speech in the transcripts. Additionally, these methods appear to be generalizable across people and languages. Speech measures extracted this way have an advantage over more commonly used clinical measures, such as neuropsychiatric tests, because they are obtained easily without the need for trained personnel. Acquisition is also efficient, fully replicable, and the data can be obtained remotely.

Despite these strengths, there are also some limitations in the method and study. f0 pitch-tracking by SAD may be inaccurate, due to voice characteristics that make it difficult to track pitch. These include vocal fry and diplophonia. Other limitations include the small number of PSPS-CBS+naPPA participants that limited statistical power. Investigation of a larger group is needed to better understand the manifestation of the combined diseases. Another issue is that we did not have sufficient motor scores to investigate possible motor effects on PSPS-CBS’s speech due to extrapyramidal impairment. We do not have an objective marker of speech errors, particularly since these are often not composed of speech sounds from the English language. Also, there was a floor effect for partial words that limited correlation analyses. Finally, longitudinal data were not available for most of our cohort.

With these limitations in mind, our findings provide a quantitative, comparative study investigating specific speech and language impairments in PSPS-CBS patients and naPPA patients. Both groups produce shorter speech segments, longer and more frequent pauses, as well as reduced f0 range and speech rate, compared to normal speakers. Patients with naPPA produced speech segments that were even shorter and pause rate even higher, and additionally show increased production of partial words and reduced verb production. Longer pauses, shorter speech segments and higher pause rate were related to dysfluency as well as impaired grammaticality in both patients with naPPA and patients with PSPS-CBS, consistent with the idea that these patient groups have speech characteristics that are on the same spectrum of impairment. This study shows how automated speech analyses can be used to characterize speech patterns in PSPS-CBS spectrum disorders.

## ACKNOWLEDGMENTS

The authors would like to thank our colleagues and staff at the Linguistic Data Consortium, Frontotemporal Degeneration Center, Penn Digital Neuropathology Laboratory, and Pathology and Laboratory Medicine Department for their contributions to our data collection and analysis, and revisions of the manuscript, as well as the caregivers and patients of this study.

## REFERENCES

[1] Armstrong MJ, Litvan I, Lang AE, Bak TH, Bhatia KP, Borroni B, Boxer AL, Dickson DW, Grossman M, Hallett M, Josephs KA, Kertesz A, Lee SE, Miller BL, Reich SG, Riley DE, Tolosa E, Troster AI, Vidailhet M, Weiner WJ (2013) Criteria for the diagnosis of corticobasal degeneration. Neurology 80, 496–503.

[2] Burrell JR, Hodges JR, Rowe JB (2014) Cognition in corticobasal syndrome and progressive supranuclear palsy: a review. Mov Disord 29, 684–693.

[3] Hoglinger GU, Respondek G, Stamelou M, Kurz C, Josephs KA, Lang AE, Mollenhauer B, Muller U, Nilsson C, Whitwell JL, Arzberger T, Englund E, Gelpi E, Giese A, Irwin DJ, Meissner WG, Pantelyat A, Rajput A, van Swieten JC, Troakes C, Antonini A, Bhatia KP, Bordelon Y, Compta Y, Corvol JC, Colosimo C, Dickson DW, Dodel R, Ferguson L, Grossman M, Kassubek J, Krismer F, Levin J, Lorenzl S, Morris HR, Nestor P, Oertel WH, Poewe W, Rabinovici G, Rowe JB, Schellenberg GD, Seppi K, van Eimeren T, Wenning GK, Boxer AL, Golbe LI, Litvan I, Movement Disorder Society-endorsed PSP Study Group (2017) Clinical diagnosis of progressive supranuclear palsy: The movement disorder society criteria. Mov Disord 32, 853–864.

[4] McMillan CT, Boyd C, Gross RG, Weinstein J, Firn K, Toledo JB, Rascovsky K, Shaw L, Wolk DA, Irwin DJ, Lee EB, Trojanowski JQ, Grossman M (2016) Multimodal imaging evidence of pathology-mediated disease distribution in corticobasal syndrome. Neurology 87, 1227–1234.

[5] Albert ML, Feldman RG, Willis AL (1974) The ‘subcortical dementia’ of progressive supranuclear palsy. J Neurol Neurosurg Psychiatry 37, 121–130.

[6] Bak TH, Crawford LM, Hearn VC, Mathuranath PS, Hodges JR (2005) Subcortical dementia revisited: similarities and differences in cognitive function between progressive supranuclear palsy (PSP), corticobasal degeneration (CBD) and multiple system atrophy (MSA). Neurocase 11, 268–273.

[7] Cotelli M, Borroni B, Manenti R, Alberici A, Calabria M, Agosti C, Arevalo A, Ginex V, Ortelli P, Binetti G, Zanetti O, Padovani A, Cappa SF (2006) Action and object naming in frontotemporal dementia, progressive supranuclear palsy, and corticobasal degeneration. Neuropsychology 20, 558–565.

[8] Graham NL, Bak T, Patterson K, Hodges JR (2003) Language function and dysfunction in corticobasal degeneration. Neurology 61, 493–499.

[9] Gurd JM, Hodges JR (1997) Word-retrieval in two cases of progressive supranuclear palsy. Behav Neurol 10, 31–41.

[10] Giannini LAA, Xie SX, McMillan CT, Liang M, Williams A, Jester C, Rascovsky K, Wolk DA, Ash S, Lee EB, Trojanowski JQ, Grossman M, Irwin DJ (2019) Divergent patterns of TDP-43 and tau pathologies in primary progressive aphasia. Ann Neurol 85, 630–643.

[11] Spinelli EG, Mandelli ML, Miller ZA, Santos-Santos MA, Wilson SM, Agosta F, Grinberg LT, Huang EJ, Trojanowski JQ, Meyer M, Henry ML, Comi G, Rabinovici G, Rosen HJ, Filippi M, Miller BL, Seeley WW, Gorno-Tempini ML (2017) Typical and atypical pathology in primary progressive aphasia variants. Ann Neurol 81, 430–443.

[12] Ash S, Moore P, Vesely L, Gunawardena D, McMillan C, Anderson C, Avants B, Grossman M (2009) Non-Fluent Speech in Frontotemporal Lobar Degeneration. J Neurolinguistics 22, 370–383.

[13] Ash S, Evans E, O’Shea J, Powers J, Boller A, Weinberg D, Haley J, McMillan C, Irwin DJ, Rascovsky K, Grossman M (2013) Differentiating primary progressive aphasias in a brief sample of connected speech. Neurology 81, 329–336.

[14] Ash S, Nevler N, Phillips J, Irwin DJ, McMillan CT, Rascovsky K, Grossman M (2019) A longitudinal study of speech production in primary progressive aphasia and behavioral variant frontotemporal dementia. Brain Lang 194, 46–57.

[15] Gorno-Tempini ML, Hillis AE, Weintraub S, Kertesz A, Mendez M, Cappa SF, Ogar JM, Rohrer JD, Black S, Boeve BF, Manes F, Dronkers NF, Vandenberghe R, Rascovsky K, Patterson K, Miller BL, Knopman DS, Hodges JR, Mesulam MM, Grossman M (2011) Classification of primary progressive aphasia and its variants. Neurology 76, 1006–1014.

[16] Grossman M, Mickanin J, Onishi K, Hughes E, D’Esposito M, Ding XS, Alavi A, Reivich M (1996) Progressive Nonfluent Aphasia: Language, Cognitive, and PET Measures Contrasted with Probable Alzheimer’s Disease. J Cogn Neurosci 8, 135–154.

[17] Gunawardena D, Ash S, McMillan C, Avants B, Gee J, Grossman M (2010) Why are patients with progressive nonfluent aphasia nonfluent? Neurology 75, 588–594.

[18] Nevler N, Ash S, Irwin DJ, Liberman M, Grossman M (2018) Validated automatic speech biomarkers in primary progressive aphasia. Ann Clin Transl Neurol 6, 4–14.

[19] Gorno-Tempini ML, Dronkers NF, Rankin KP, Ogar JM, Phengrasamy L, Rosen HJ, Johnson JK, Weiner MW, Miller BL (2004) Cognition and anatomy in three variants of primary progressive aphasia. Ann Neurol 55, 335–346.

[20] Boeve B, Dickson D, Duffy J, Bartleson J, Trenerry M, Petersen R (2003) Progressive nonfluent aphasia and subsequent aphasic dementia associated with atypical progressive supranuclear palsy pathology. Eur Neurol 49, 72–78.

[21] Josephs KA, Boeve BF, Duffy JR, Smith GE, Knopman DS, Parisi JE, Petersen RC, Dickson DW (2005) Atypical progressive supranuclear palsy underlying progressive apraxia of speech and nonfluent aphasia. Neurocase 11, 283–296.

[22] Karnik NS, D’Apuzzo M, Greicius M (2006) Non-fluent progressive aphasia, depression, and OCD in a woman with progressive supranuclear palsy: neuroanatomical and neuropathological correlations. Neurocase 12, 332–338.

[23] Kertesz A, Martinez-Lage P, Davidson W, Munoz DG (2000) The corticobasal degeneration syndrome overlaps progressive aphasia and frontotemporal dementia. Neurology 55, 1368–1375.

[24] Lebrun Y, Devreux F, Rousseau JJ (1986) Language and speech in a patient with a clinical diagnosis of progressive supranuclear palsy. Brain Lang 27, 247–256.

[25] McMonagle P, Blair M, Kertesz A (2006) Corticobasal degeneration and progressive aphasia. Neurology 67, 1444–1451.

[26] Mimura M, Oda T, Tsuchiya K, Kato M, Ikeda K, Hori K, Kashima H (2001) Corticobasal degeneration presenting with nonfluent primary progressive aphasia: a clinicopathological study. J Neurol Sci 183, 19–26.

[27] Mochizuki A, Ueda Y, Komatsuzaki Y, Tsuchiya K, Arai T, Shoji S (2003) Progressive supranuclear palsy presenting with primary progressive aphasia--clinicopathological report of an autopsy case. Acta Neuropathol 105, 610–614.

[28] Murray R, Neumann M, Forman MS, Farmer J, Massimo L, Rice A, Miller BL, Johnson JK, Clark CM, Hurtig HI, Gorno-Tempini ML, Lee VM, Trojanowski JQ, Grossman M (2007) Cognitive and motor assessment in autopsy-proven corticobasal degeneration. Neurology 68, 1274–1283.

[29] Takao M, Tsuchiya K, Mimura M, Momoshima S, Kondo H, Akiyama H, Suzuki N, Mihara B, Takagi Y, Koto A (2006) Corticobasal degeneration as cause of progressive non-fluent aphasia: clinical, radiological and pathological study of an autopsy case. Neuropathology 26, 569–578.

[30] Rohrer JD, Paviour D, Bronstein AM, O’Sullivan SS, Lees A, Warren JD (2010) Progressive supranuclear palsy syndrome presenting as progressive nonfluent aphasia: a neuropsychological and neuroimaging analysis. Mov Disord 25, 179–188.

[31] Santos-Santos MA, Mandelli ML, Binney RJ, Ogar J, Wilson SM, Henry ML, Hubbard HI, Meese M, Attygalle S, Rosenberg L, Pakvasa M, Trojanowski JQ, Grinberg LT, Rosen H, Boxer AL, Miller BL, Seeley WW, Gorno-Tempini ML (2016) Features of Patients With Nonfluent/Agrammatic Primary Progressive Aphasia With Underlying Progressive Supranuclear Palsy Pathology or Corticobasal Degeneration. JAMA Neurol 73, 733–742.

[32] Mathew R, Bak TH, Hodges JR (2011) Screening for cognitive dysfunction in corticobasal syndrome: utility of Addenbrooke’s cognitive examination. Dement Geriatr Cogn Disord 31, 254–258.

[33] Irwin DJ, Lleo A, Xie SX, McMillan CT, Wolk DA, Lee EB, Van Deerlin VM, Shaw LM, Trojanowski JQ, Grossman M (2017) Ante mortem cerebrospinal fluid tau levels correlate with postmortem tau pathology in frontotemporal lobar degeneration. Ann Neurol 82, 247–258.

[34] Golbe LI, Ohman-Strickland PA (2007) A clinical rating scale for progressive supranuclear palsy. Brain 130, 1552–1565.

[35] Cho S, Nevler N, Ash S, Shellikeri S, Irwin DJ, Massimo L, Rascovsky K, Olm C, Grossman M, and Liberman M (2020) Automated analysis of lexical features in frontotemporal degeneration. medRxiv. DOI: https://doi.org/10.1101/2020.09.10.20192054.

[36] Nevler N, Ash S, Jester C, Irwin DJ, Liberman M, Grossman M (2017) Automatic measurement of prosody in behavioral variant FTD. Neurology 89, 650–656.

[37] Nevler N, Ash S, McMillan C, Elman L, McCluskey L, Irwin DJ, Cho S, Liberman M, Grossman M (2020) Automated Analysis of Natural Speech in Amyotrophic Lateral Sclerosis Spectrum Disorders. Neurology, 10.1212/WNL.0000000000010366.

[38] Goodglass H, Kaplan E (1972) Boston diagnostic aphasia examination (BDAE), Lea & Febiger, Philadelphia.

[39] Ryant, N (2013) LDC HMM Speech Activity Detector, v.1.0.4, Linguistic Data Consortium, University of Pennsylvania.

[40] Yuan J, Neville R, Liberman M, Stolcke A, Mitra V, and Wang W (2013) Automatic phonetic segmentation using boundary models. In Proceedings of the Annual Conference of the International Speech Communication Association, INTERSPEECH, 2306–2310.

[41] Boersma P, Weenink D 1992-2014), Praat, v.5.3.76, Institute of Phonetic Sciences, University of Amsterdam.

[42] Cho S, Nevler N, Shelliker S, Parjane N, Irwin DJ, Neville R, Ash S, Cieri C, Liberman M, Grossman M (2020) Lexical and acoustic characteristics of young and older healthy adults. Accepted for publication in JSLHR.

[43] Honnibal M, Johnson M (2015) An improved non-monotonic transition system for dependency parsing. In Proceedings of the 2015 conference on empirical methods in natural language processing, 1373–1378.

[44] Industrial-Strength Natural Language Processing: IN PYTHON, https://spacy.io/, Last updated 2020, Accessed in 2019-2020.

[45] Marcus MP, Santorini B, Marcinkiewicz MA (1993). Building a Large Annotated Corpus of English: The Penn Treebank. Computational Linguistics, 19, 313–330.

[46] Petrov S, Das D, McDonald R (2012). A Universal Part-of-Speech Tagset. Proceedings of the Eighth International Conference on Language Resources and Evaluation, European Language Resources Association, 2089–2096.

[47] RStudio Team (2019). RStudio: Integrated Development for R. RStudio, Inc., Boston, MA URL http://www.rstudio.com/.

[48] Kazuki Yoshida (2020). tableone: Create ‘Table 1’ to Describe Baseline Characteristics. R package version 0.11.1. https://CRAN.R-project.org/package=tableone.

[49] Dag, O., Dolgun, A., Konar, N.M. (2018). onewaytests: An R Package for One-Way Tests in Independent Groups Designs. The R Journal, 10:1, 175–199.

[50] Peters G (2018). _userfriendlyscience: Quantitative analysis made accessible_. DOI: 10.17605/osf.io/txequ (URL: https://doi.org/10.17605/osf.io/txequ), R package version 0.7.2, URL : https://userfriendlyscience.com.

[51] Hadley Wickham (2007). Reshaping Data with the reshape Package. Journal of Statistical Software, 21(12), 1–20. URL http://www.jstatsoft.org/v21/i12/.

[52] Manuel Morales, with code developed by the R Development Core Team, with general advice from the R-help listserv community and especially Duncan Murdoch. (2020). sciplot: Scientific Graphing Functions for Factorial Designs. R package version 1.2-0. https://CRAN.R-project.org/package=sciplot.

[53] Frank E Harrell Jr, with contributions from Charles Dupont and many others. (2020). Hmisc: Harrell Miscellaneous. R package version 4.4-0. https://CRAN.R-project.org/package=Hmisc.

[54] Rene Locher (2020). IDPmisc: ‘Utilities of Institute of Data Analyses and Process Design (www.zhaw.ch/idp)’. R package version 1.1.20. https://CRAN.R-project.org/package=IDPmisc.

[55] Josephs KA, Duffy JR (2008) Apraxia of speech and nonfluent aphasia: a new clinical marker for corticobasal degeneration and progressive supranuclear palsy. Curr Opin Neurol 21, 688–692.

[56] Sitek EJ, Kluj-Kozłowska K, Barczak A, Kozłowski M, Wieczorek D, Przewłócka A, Narożańska E, Dąbrowska M, Barcikowska M, Sławek J (2015) Overlapping and distinguishing features of descriptive speech in Richardson variant of progressive supranuclear palsy and non-fluent progressive aphasia. Postępy Psychiatrii i Neurologii 24, 62–67.

[57] Skodda S, Visser W, Schlegel U (2011) Acoustical analysis of speech in progressive supranuclear palsy. J Voice 25, 725–731.

[58] Strand EA, Duffy JR, Clark HM, Josephs K (2014) The Apraxia of Speech Rating Scale: a tool for diagnosis and description of apraxia of speech. J Commun Disord 51, 43–50.

